# Development and validation of a digital pathology artificial intelligence (DPAI)-based biomarker predicting risk of Gleason grade group reclassification for patients who are candidates for active surveillance

**DOI:** 10.64898/2026.05.15.26353328

**Authors:** Brent Mabey, Lauren H. Lenz, Matthew J. Schiewer, Walter Rayford, Hassan Muhammad, Wei Huang, Robert Finch, Christina Nakamoto, Hosein Kouros-Mehr, Jeff Jasper, Hirak Basu, Chao Feng, Anurag Sharma, George Wilding, Rajat Roy, Dale Muzzey, Alexander Gutin

**Affiliations:** Myriad Genetics, Inc., Salt Lake City, Utah, USA; The Urology Group, Memphis, TN, USA; PATHOMIQ, Inc., Cupertino, CA, USA; Department of Pathology and Laboratory Medicine, University of Wisconsin-Madison, Madison, WI, USA

**Keywords:** prostate cancer, active surveillance, grade reclassification, digital pathology artificial intelligence, predictive biomarker

## Abstract

**Aims:** Active surveillance (AS) allows selected men with localized prostate cancer to defer curative therapy and reduce treatment morbidity. Conversion from AS to treatment is commonly triggered by Gleason grade group (GGG) upgrading on confirmatory biopsy. We developed and validated a digital pathology artificial intelligence (DPAI) biomarker to predict GGG upgrading in AS-eligible patients.

**Materials & Methods:** The DPAI model was trained using histopathology image features from diagnostic biopsies of 998 patients and validated in an independent cohort of 296 patients meeting criteria for AS. Logistic regression estimated the probability of confirmatory-biopsy GGG increase, and feature selection identified the most predictive variables.

**Results:** AI-GUR (Artificial Intelligence-Gleason Upgrade Risk) predicted GGG reclassification at confirmatory biopsy (OR 1.60; p=0.0003), and provided information beyond conventional stratification (risk group, CAPRA) and cribriform morphology (all p<0.01). Predicted risks were similar across time from diagnosis (∼10-15% to ∼85% at 1, 1.5, or 2 years; p for time=0.50), consistent with initial biopsy mischaracterization rather than time-dependent progression.

**Conclusions:** AI-GUR provides individualized estimates of confirmatory-biopsy GGG upgrading for AS candidates. Using DPAI may improve shared decision-making by complementing standard clinicopathologic tools and molecular testing using the same biopsy specimen, while informing the likelihood of grade upgrade at confirmation.

**Summary Points:** Active surveillance (AS) can reduce treatment-related morbidity in localized prostate cancer (PCa), and confirmatory biopsy is used to address uncertainty related to grade reclassification and potential mischaracterization at diagnostic biopsy.

We developed and validated AI-GUR (Artificial Intelligence–Gleason Upgrade Risk), a digital pathology AI (DPAI) biomarker to predict Gleason grade group (GGG) reclassification at confirmatory biopsy in AS-eligible patients.

The model was trained in 998 patients and validated in an independent cohort of 296 patients, all candidates for AS by guideline criteria.

In validation, AI-GUR significantly predicted GGG reclassification (OR 1.60, p=0.0003) and added information beyond conventional risk stratification tools (risk group, CAPRA) and cribriform morphology (p-values <0.01).

Predicted risk was invariant with time since diagnosis (∼10–85% at 1, 1.5, or 2 years; p-value for time=0.50), consistent with capturing mischaracterization at initial biopsy rather than time-dependent progression; AI-GUR provides individualized risk estimates that may improve shared decision-making for AS candidates using the same biopsy material used for conventional stratification and advanced molecular prognostic testing, with a limitation of archived material.

## Introduction

Active surveillance (AS) allows patients with appropriately selected prostate cancer (PCa) risk profiles to safely avoid/delay morbidity associated with definitive therapy by closely monitoring disease and intervening upon progression. Despite strong long-term evidence supporting safety, AS remains underutilized in the United States, with only up to 60% of eligible patients electing this management strategy[1]. This gap persists largely due to uncertainty at the earliest decision points of care, where fear of undertreatment or missed opportunity for cure outweighs the objectively low risk of metastasis and PCa-specific mortality in patients with Gleason grade group 1 (GGG1) and select favorable intermediate-risk disease[2]. Tools that improve individualized risk assessment and support shared decision-making across the key phases of AS—candidacy, confirmation of candidacy, and ongoing surveillance—have the potential to increase appropriate AS utilization while improving patient confidence in surveillance-based management.

Identifying patients for AS candidacy largely relies on pathological assessment of diagnostic biopsies. National Comprehensive Cancer Network® (NCCN®) Clinical Practice Guidelines in Oncology®[2] state that AS is the preferred management strategy for GGG1/low-risk disease and AS should be considered for GGG2/favorable intermediate-risk disease without unfavorable histology. Confirmation of AS candidacy includes pathological assessment of a follow-up biopsy, which allows for early detection of grade reclassification or cancer progression and mitigates any underestimates of tumor grade in the diagnostic biopsy due to baseline tumor heterogeneity, sampling variability, and subjective grading systems[2]. The following pathological considerations have been enumerated as rationale for transitioning from AS to definitive therapy: reclassification from GGG1 to ≥GGG2 in patients with low-risk disease; grade reclassification from GGG2 to higher, increase in percent Gleason Pattern 4 (GP4), increase in tumor volume, or rise in PSA density for patients with intermediate risk-disease[2].

Some variables captured coincident with diagnostic biopsy have been identified as prognostic for grade reclassification on confirmatory biopsies, as described in the NCCN Guidelines®: tumor burden indicators (increased number positive cores, PSA density ≥0.15, faster PSA kinetics), tumor biology (increase in percentage GP4, presence of cribriform and/or intraductal carcinoma), and host/systemic factors (such as BMI)[2]. Importantly, existence of any one ofa these features alone is insufficient to require immediate use of definitive therapy but should be considered as part of a set of factors that informs the shared decision-making process in personalizing the intensity of an AS management protocol. While it is strongly recommended that all AS candidates undergo a confirmatory biopsy, the suggested timing ranges from 6-36 months, depending on a patient’s risk profile and multi-parametric MRI findings[2].

Further refinement of risk can be provided by advanced tools, such as the clinically validated Prolaris Prognostic Test (Myriad Genetics, Inc.) which identifies patients at higher risk of poor outcomes (metastasis and PCa mortality) from diagnostic biopsy material, even in patients with GGG1 and GGG2 disease[3]. While Prolaris refines baseline risk of adverse outcomes, it does not address the probability of grade reclassification on confirmatory biopsy, which is an orthogonal but clinically critical question. Current approaches don’t allow for simultaneous refinement of initial risk as well as estimation of grade reclassification risk on confirmatory biopsy, limiting their utility in confirming AS candidacy.

Here, we describe the development and validation of a novel biomarker designed to predict grade reclassification on confirmatory biopsy in patients with localized PCa who are candidates for AS. By integrating standard-of-care NCCN risk stratification and Prolaris testing with a digital pathology artificial intelligence (DPAI) model trained on diagnostic biopsy material, this approach enables simultaneous assessment of AS candidacy and reclassification risk from a single biopsy specimen. This unified framework addresses a critical unmet need during AS confirmation, with the potential to inform the urgency of confirmatory biopsy, decrease uncertainty at early decision points, and support more confident, personalized shared decision-making. This strategy may help reduce unnecessary definitive treatment and promote more appropriate and durable implementation of AS in clinical practice.

## Materials and Methods

### PATHOMIQ AI Platform

PATHOMIQ_PRAD is an AI-powered pipeline designed to generate AI-derived risk score that predicts biochemical recurrence and metastasis in localized PCa[4-6]. It comprises multiple modules including slide quality assessment, cancer detection, and cancer grading. The pipeline also produces detailed quantification of standard histopathologic features used in PCa risk assessment, including glandular-level Gleason grade percentages and ratios, tumor volume, and other metrics.

Whole slide images of hematoxylin- and eosin-stained (H&E) slides were obtained via brightfield microscopy at 40x using the Aperio GT 450 digital slide scanner (Leica Biosystems). Images were managed with the Aperio eSLide Manager WebViewer (Leica Biosystems). The PRAD algorithm was applied to images within the HALO AP digital pathology platform (Indica Labs) in two steps as described in the Supplement.

### Histopathological Tissue Specimens

Diagnostic biopsy samples were obtained during the routine execution of commercial laboratory testing. The activities of the current report do not constitute human subjects research under 45 CFR §46 because it involved only de-identified samples, with no identifiable private information and no interaction or intervention with individuals. As a result, the project is not subject to human subjects’ research requirements, and informed consent was not required.

### Training the AI-GUR Score

The AI Gleason Upgrade Risk (AI-GUR) score was trained on two observational cohorts, as well as a cohort of patients who had biopsies taken as part of their standard clinical care and had their diagnostic biopsy and follow-up biopsy sent for Prolaris testing. Training cohorts were restricted to patients considered candidates for AS by clinical features (diagnostic GGG1 or 2). The clinical cohort was enriched to have equal numbers of GGG1 and GGG2 patients, as well as equal numbers of events and non-events. Events were defined as any increase in GGG (from GGG1 to ≥GGG2, and GGG2 to ≥GGG3). A summary of the full training cohort (n=998) is given in Supplemental Table 1, with a patient flow diagram shown in Supplemental Figure 1.

All training and validation analyses were performed in R version 4 or higher (R Foundation for Statistical Computing, 2026). AI-GUR was built by modeling the probability of GGG increase using logistic regression. Despite the timing between biopsies playing a role in GGG increase, logistic regression was used instead of survival analyses to account for the possibility of non-time-dependent factors in the events, such as resampling error and grading discordance by pathologist(s). Candidate variables for the model includes 16-dimensional AI-embedding feature and AI-assessed histopathologic features produced by the PATHOMIQ_PRAD algorithm, the PRAD score (that combines the 16-dimensional AI-embedding and AI-assessed histopathologic features[4-6]), the molecular cell cycle progression score (CCP)[7] produced by the Prolaris test, GGG at diagnosis (GGG1 vs GGG2), and interactions between diagnostic GGG and all other variables. AI-assessed histopathologic features measured as a proportion were logit transformed. Stepwise feature selection was used to choose candidate variables, starting from a model with all features included, then adding or dropping variables at each step based on their significance in a Chi-squared likelihood ratio test, with a cutoff at p=0.05.

After feature selection, the final model included two AI-assessed histopathologic features (proportion of identified cancer area with GP4, cribriform morphology, or perineural invasion, and the proportion of tissue area identified as tumor) as well as diagnostic Gleason and an interaction between diagnostic Gleason and proportion of tissue area identified as tumor.

The validation analysis set remained blinded until after the final model was locked.

### Statistical methods for validating the AI-GUR Score

Analyses followed a pre-defined statistical analysis plan, with non-pre-specified analyses labeled exploratory. Inclusion criteria were pre-specified and included requiring diagnostic GGG1 or GGG2, being identified as a candidate for active surveillance by the Prolaris test (CCR score ≤ 0.8[8]), known time to confirmatory biopsy, and GGG of confirmatory biopsy. Patients for whom a diagnostic biopsy slide was unavailable, or an AI-GUR score could not be calculated were excluded from the analysis set (Supplemental Figure 1).

The endpoint of interest was any increase in GGG at confirmatory biopsy, referred to as GGG reclassification.

Distributions of continuous variables were compared graphically, as well as with exploratory Kolmogorov-Smirnov tests and correlations. Rates were compared with tests of two proportions. The relationship between the AI-GUR score and other variables of interest and GGG upgrade status was evaluated using logistic regression models, accounting for cohort as a random effect. Results are reported as odds ratios (OR) per standard deviation (SD) for continuous variables, with 95% profile-likelihood confidence intervals and likelihood ratio test p-values. Significance was pre-defined at the alpha = 0.05 level.

Distributions of the AI-GUR score and associated risks of GGG reclassification were examined separately for GGG1 and GGG2 patients in a set of samples comprising the observational studies used in training and validation as well as a group of commercially tested patients (Supplemental Figure 1). These distributions were used to calculate the 20^th^ and 80^th^ percentile of scores within each GGG group.

## Results

### Validation Cohort Characteristics

The validation cohort included N = 296 patients from two prospective, observational studies and one retrospective, observational study (Supplemental Figure 1). The retrospective and one of the prospective studies were randomly split between training and validation (Supplemental Figure 1). A substantial majority of patients were NCCN low risk (81.8%) and GGG1 (84.8%). One third (99/296, 33.4%) of patients experienced GGG reclassification at confirmatory biopsy (i.e., the first biopsy following the diagnostic biopsy). The median time from diagnosis to confirmatory biopsy was 1.2 years (interquartile range 0.8 to 1.6 years; maximum 5.0 years; Supplemental Figure 2). A summary of the validation cohort is given in Table 1, including self-reported ancestry.

**Table 1.**
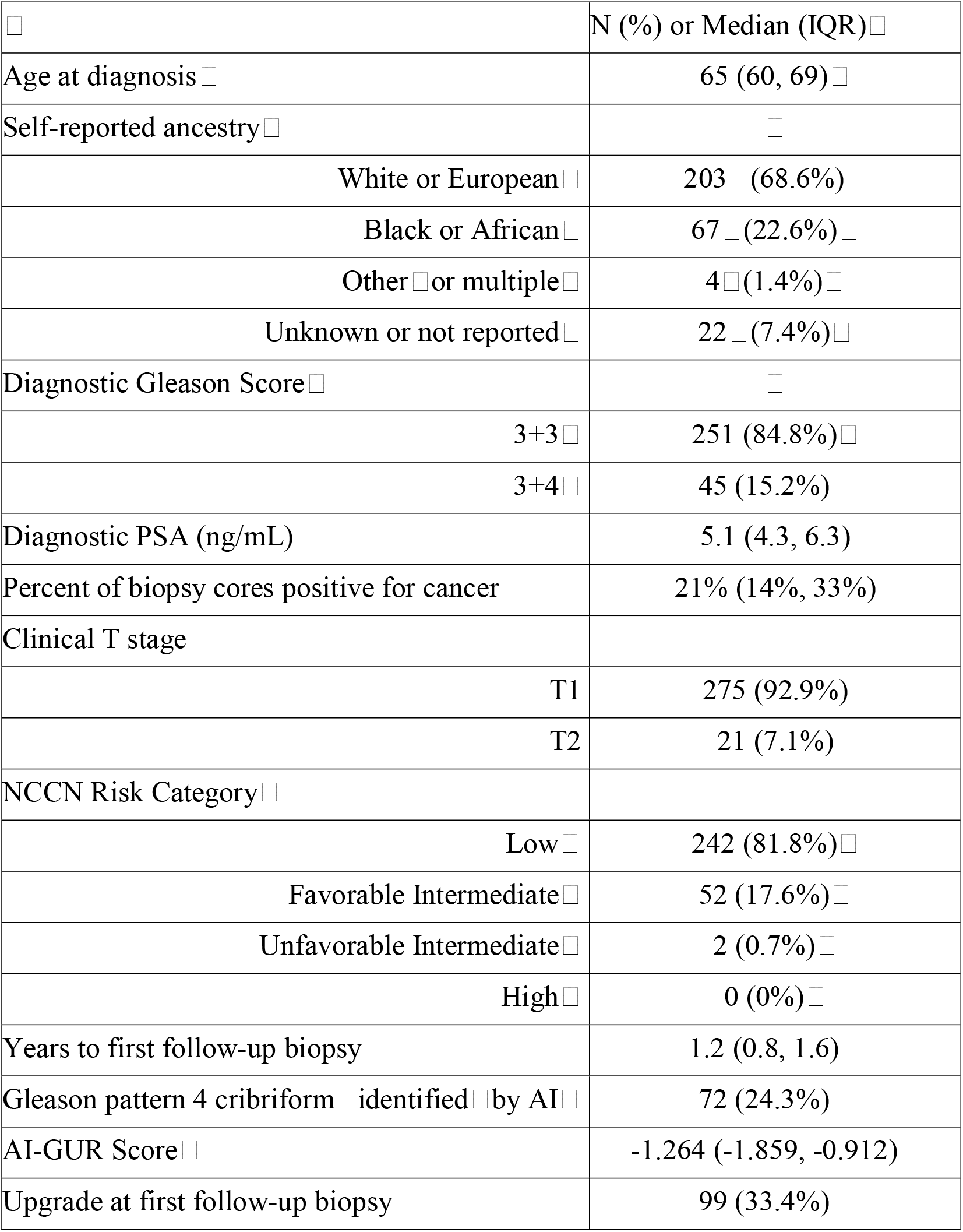
□Characteristics of the validation cohort, N = 296. □ □.

|  | N (%) or Median (IQR) |
| --- | --- |
| Age at diagnosis | 65 (60, 69) |
| Self-reported ancestry |  |
| White or European | 203 (68.6%) |
| Black or African | 67 (22.6%) |
| Other or multiple | 4 (1.4%) |
| Unknown or not reported | 22 (7.4%) |
| Diagnostic Gleason Score |  |
| 3+3 | 251 (84.8%) |
| 3+4 | 45 (15.2%) |
| Diagnostic PSA (ng/mL) | 5.1 (4.3, 6.3) |
| Percent of biopsy cores positive for cancer | 21% (14%, 33%) |
| Clinical T stage |  |
| T1 | 275 (92.9%) |
| T2 | 21 (7.1%) |
| NCCN Risk Category |  |
| Low | 242 (81.8%) |
| Favorable Intermediate | 52 (17.6%) |
| Unfavorable Intermediate | 2 (0.7%) |
| High | 0 (0%) |
| Years to first follow-up biopsy | 1.2 (0.8, 1.6) |
| Gleason pattern 4 cribriform identified by AI | 72 (24.3%) |
| AI-GUR Score | -1.264 (-1.859, -0.912) |
| Upgrade at first follow-up biopsy | 99 (33.4%) |

### Validation of DPAI Gleason Upgrade Predictive Score (AI-GUR)

In a model with AI-GUR score and time between diagnosis and confirmatory biopsy predicting GGG upgrade at confirmatory biopsy, AI-GUR score was a statistically significant predictor (OR 1.60, 95% CI 1.23-2.11, p = 0.0003), whereas time (in years) since diagnosis was not (OR 1.09, 95% CI 0.85-1.38, p = 0.50; Table 2). The probability of GGG reclassification at first follow-up biopsy vs. AI-GUR score is shown in Figure 1, which demonstrates that with increasing AI-GUR score the probability of GGG reclassification increases similarly (from ∼10% to ∼85%) irrespective of time elapsed between biopsies (1, 1.5, or 2 years).

**Figure 1:**
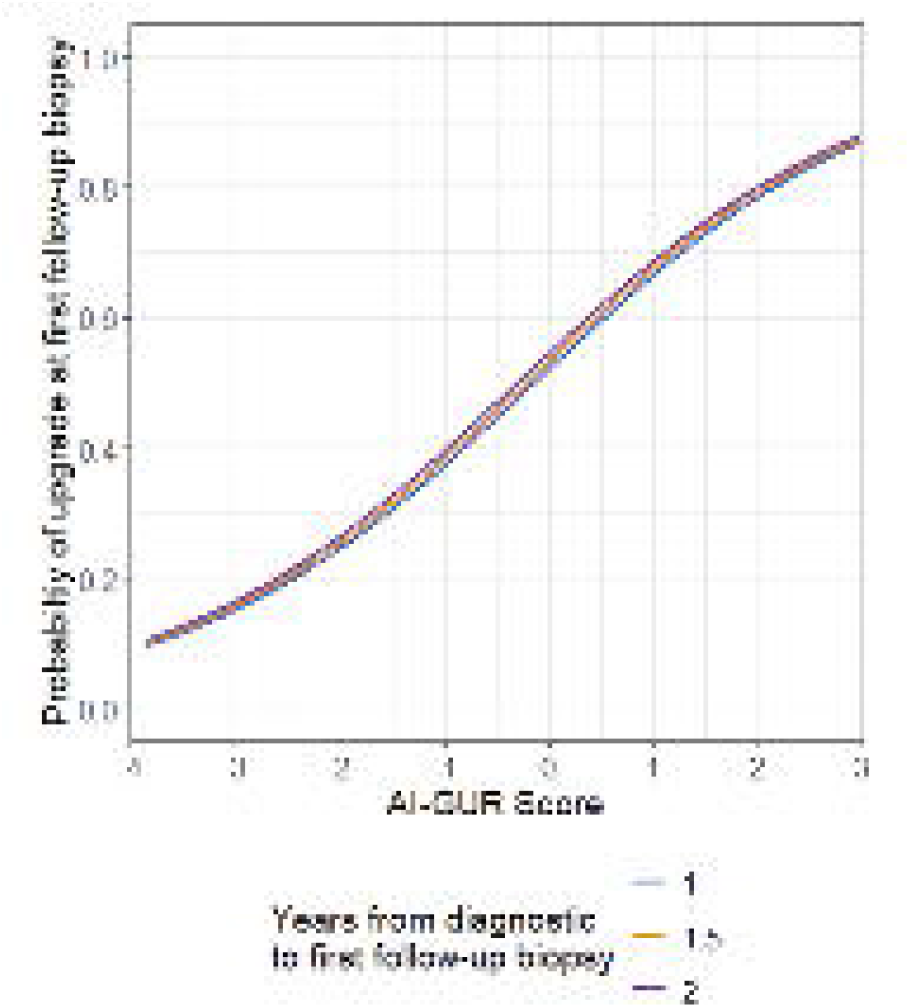
Probability of Gleason upgrade at first follow-up biopsy vs AI-GUR for follow-up biopsies at 1, 1.5, and 2 years post-diagnosis.

**Table 2.** □Results of□ a□ multivariate logistic regression model□ predicting Gleason upgrade at first follow-up biopsy.□ Model includes□ a random effect for study. □ □.

| Predictor | Odds Ratio per standard deviation (95% confidence interval) | P-value |
| --- | --- | --- |
| AI-GUR | 1.60 (1.23, 2.11) | 0.0003 |
| Years since diagnosis | 1.09 (0.85, 1.38) | 0.50 |

After accounting for diagnostic GGG, AI-GUR score remained significantly associated with risk of GGG reclassification (p=0.02), while years since diagnosis and diagnostic GGG were not statistically significant (p=0.53 and p=0.33, respectively; Table 3A). This indicates that the AI-based portion of the AI-GUR score contributes significant information about the risk of GGG reclassification above and beyond the information contained in the diagnostic GGG.

**Table 3.** □Results of multivariable logistic regression models with AI-GUR, □ years between diagnosis and first follow-up biopsy, and clinical variables predicting Gleason upgrade.□ Models include a random effect□ for □ study. □ □.

| Predictor | Odds Ratio per standard deviation (95% confidence interval) | P-value |
| --- | --- | --- |
| AI-GUR | 1.46 (1.06, 2.03) | 0.02 |
| Years since diagnosis | 1.08 (0.85, 1.37) | 0.53 |
| Diagnostic Gleason Grade Group |  | 0.33 |
| GGG1 | Reference |  |
| GGG2 | 0.61 (0.21, 1.62) |  |

B: AI-GUR, years between diagnosis and first follow-up biopsy, and log(diagnostic PSA+1) predicting Gleason upgrade.
| Predictor | Odds Ratio per standard deviation (95% confidence interval) | P-value |
| --- | --- | --- |
| AI-GUR | 1.60 (1.23, 2.11) | $3.2 \times 10^{-4}$ |
| Years since diagnosis | 1.12 (0.88, 1.43) | 0.35 |
| Log(diagnostic PSA +1) | 1.30 (1.01, 1.68) | 0.04 |

In exploratory analyses, AI-GUR also added significant predictive information to other clinical variables, including CAPRA score, NCCN risk category, and their components (all p-values < 0.01, not shown). The only clinical feature that added significant predictive information to AI-GUR was diagnostic PSA (transformed as log(PSA +1)), which was only slightly significant (p = 0.04, Table 3B). This indicates that the AI-GUR score provides significant, independent information about risk of Gleason upgrade above and beyond the information contained in clinical variables.

Additionally, no meaningful correlation was observed between AI-GUR and the Prolaris score (CCR) (r=-0.18) or its components (CCP and CAPRA) (r=-0.03 and r=-0.22, respectively) (Figure 2A-C), indicating the AI-GUR score contains information independent of these variables.

**Figure 2:**
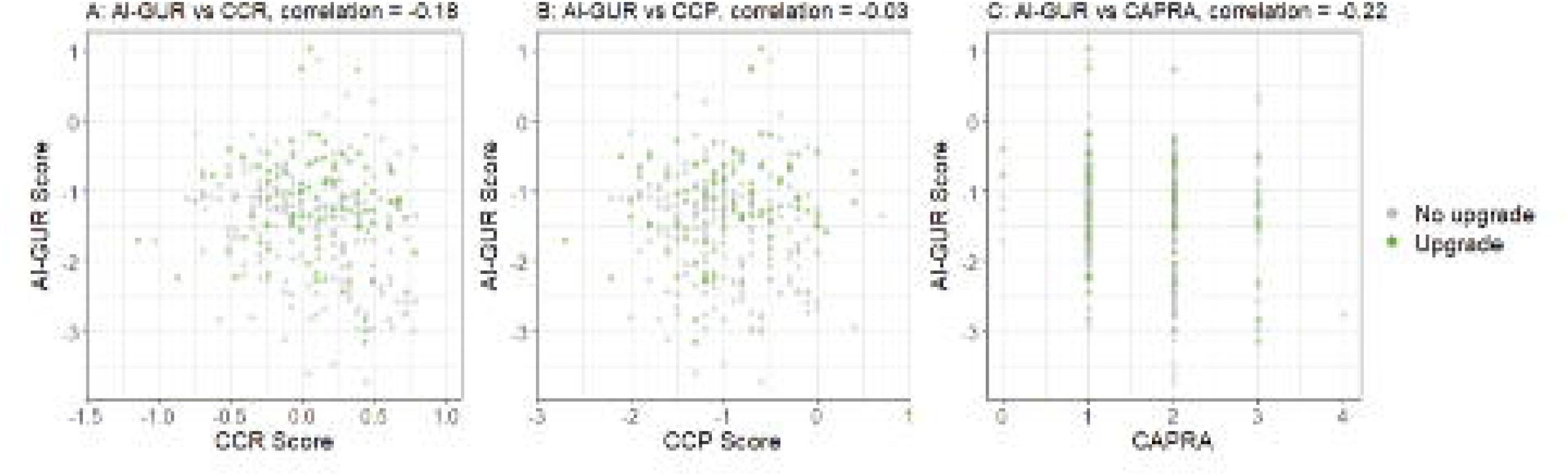
Distributions of Al-GUR compared to (A) the Prolaris combined clinical risk (CCR) score or its components, (B) the cell-cycle progression (CCP) score and (C) the University of California San Francisco Cancer of the Prostate Risk Assessment (CAPRA) score.

As part of the PATHOMIQ_PRAD algorithm, cribriform tissue can be reported and analyzed as a binary (present yes or no), as a proportion of total tissue area, or as a proportion of area of cancer tissue. In the validation cohort, AI-identified GP4 cribriform morphology was present in 22.8% (45 of 197) of patients who did not have a GGG reclassification event and in 27.3% (27 of 99) patients with a GGG reclassification event (p=0.49). Where GP4 cribriform was identified, the extent of cribriform ranged from 0.01% to 37.9% of total AI-identified tissue area, and 0.05% to 60.7% of total AI-identified cancer tissue area. The distribution of extent of cribriform morphology by GGG reclassification status is shown in Supplemental Figure 3. As shown in Table 4A-C, the presence (binary yes/no) or extent of GP4 cribriform (as proportion of total tissue area or tumor tissue area) did not add significant information to the AI-GUR score when predicting GGG upgrade at confirmatory biopsy (all p-values >0.1), whereas AI-GUR score remained significant (all p-values <0.001). These data indicate that the AI-GUR score, which contains information about cribriform morphology, contains more predictive information than does cribriform morphology alone.

**Table 4.**
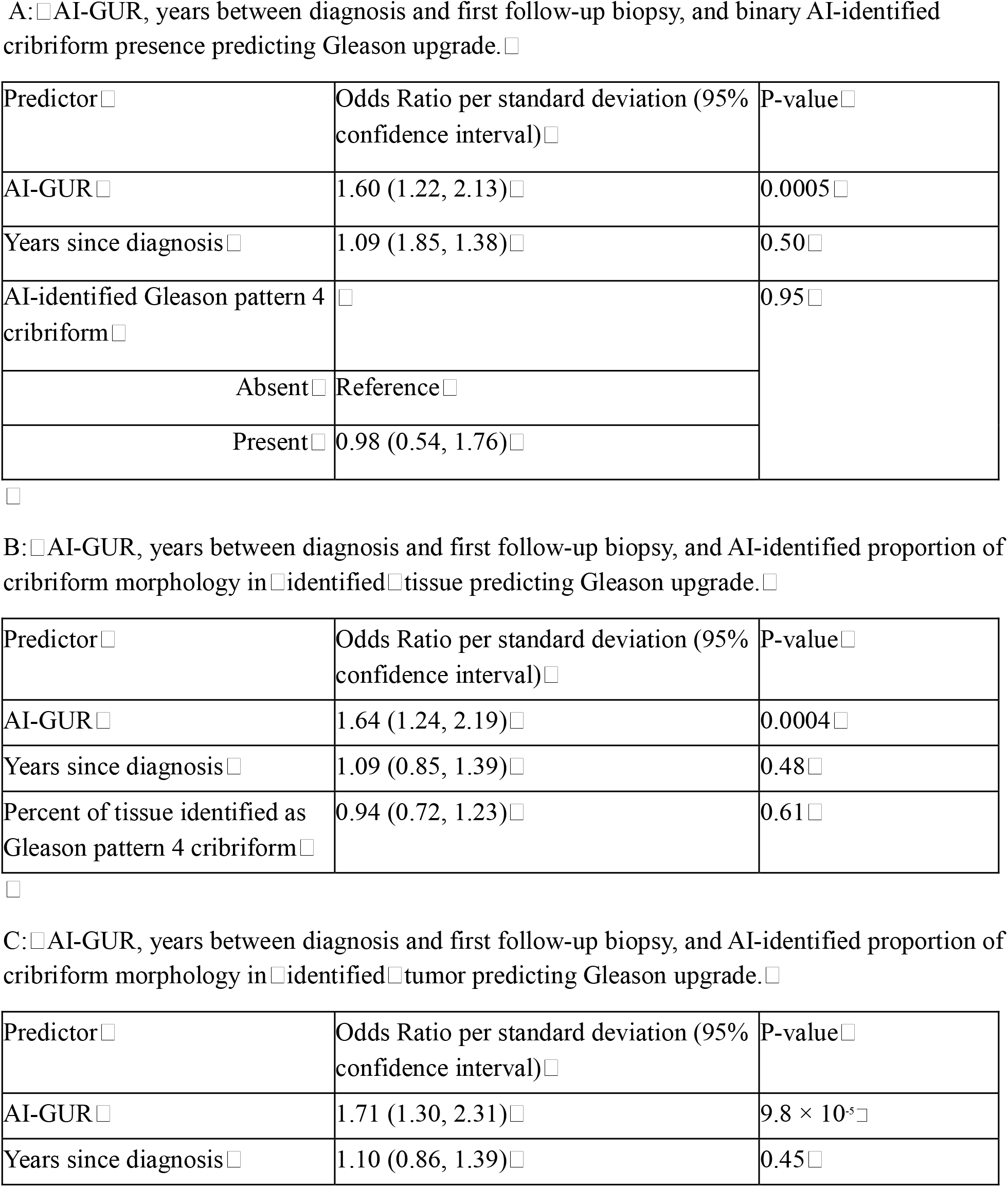
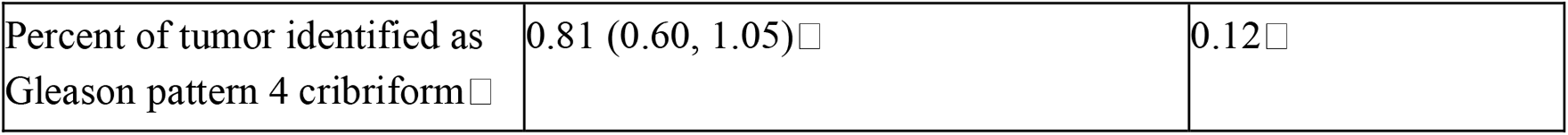
□Results of multivariate logistic regression models with AIGUR, years between diagnosis and first follow-up biopsy, and AI-identified cribriform variables □ predicting Gleason upgrade at first follow-up biopsy. Models □ includes □ a random effect for study.□ □.

### Distributions of AI-GUR

The distributions of AI-GUR scores in N = 916 GGG1 patients and N = 478 GGG2 patients clinically tested with Prolaris and identified as candidates for AS by Prolaris are shown in Figure 3. The distributions differed by diagnostic GGG, indicating that GGG2 patients were less likely to experience GGG reclassification than GGG1 patients. In GGG1 patients, the 20^th^ and 80^th^ percentiles of AI-GUR scores were −1.646 and −0.767, respectively, corresponding to risks of GGG reclassification of 29.64% and 41.75% based on data observed in the validation cohort. The 20^th^ and 80^th^ percentiles of AI-GUR scores in GGG2 patients were −2.885 and −2.151, respectively, corresponding to risks of 16.62% and 23.70%.

**Figure 3:**
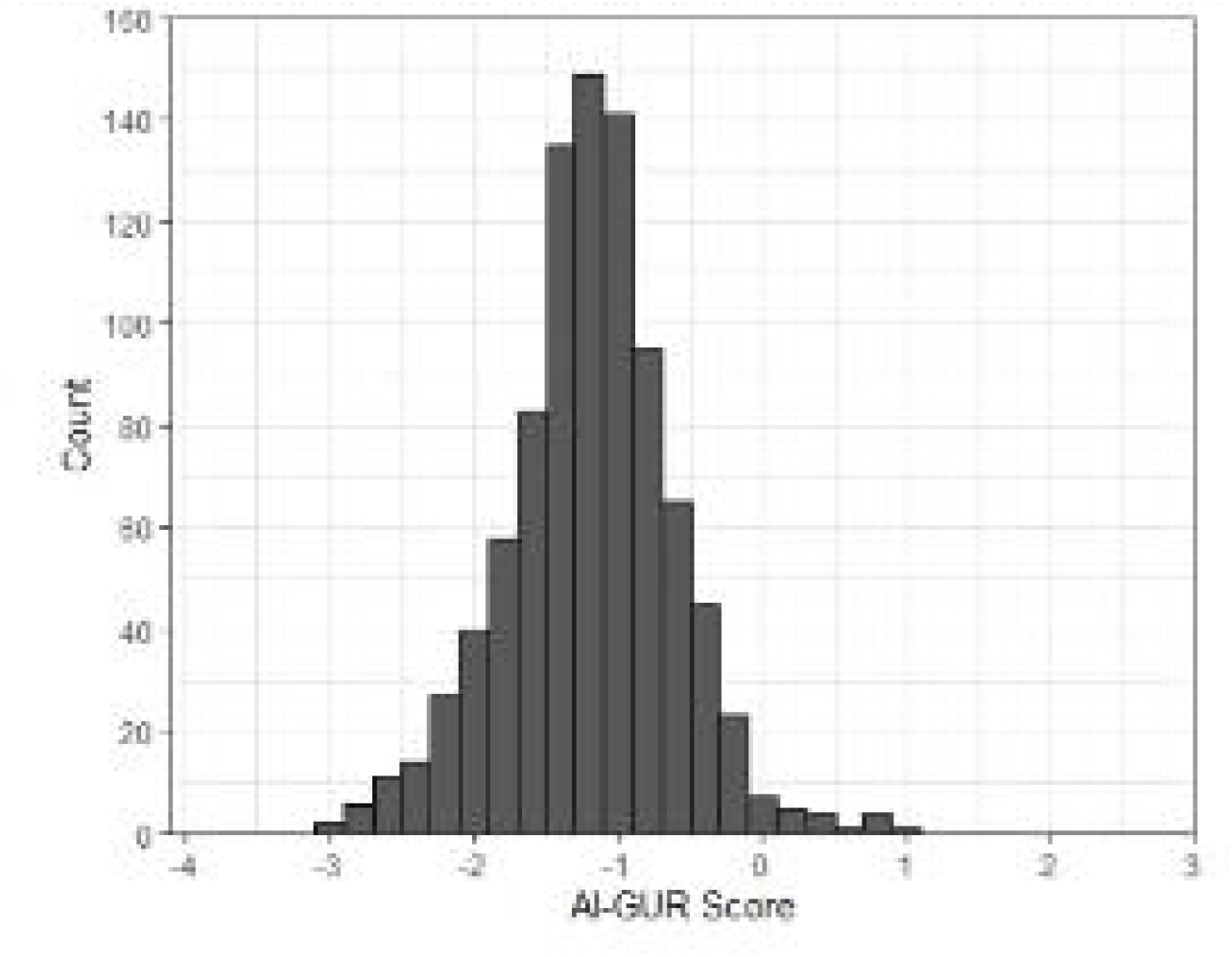
AI-GUR scores in the distribution analysis set, separated by diagnostic Gleason grade. A. Diagnostic Gleason 3+3, N-916. Median-1.181, interquartile range -1.532 to -0.857.

## Discussion

We developed and validated a novel diagnostic biopsy DPAI model that predicts GGG reclassification events on confirmatory biopsies for patients who are candidates for AS. This model was trained on diagnostic biopsy tissues from patients who were identified as candidates for AS by conventional risk stratification. Validation was conducted in biopsy tissues from patients who were identified as candidates for AS by conventional risk stratification as well as by molecular testing (Prolaris, CCR). Importantly, AI-GUR may address an unmet need described in the NCCN Guidelines, i.e., that there is not an advanced risk stratification tool with sufficient evidence to identify patients at elevated risk of GGG reclassification during the AS candidacy confirmation process[2]. To our knowledge, the AI-GUR model described herein is the first AI-powered H&E image-based morphometric biomarker with validity at this critical decision point.

DPAI-based prognostic and predictive models must demonstrate added value beyond conventional pathological features routinely used in clinical decision-making, including GGG and cribriform morphology. In the present study, AI-GUR remained independently predictive of GGG reclassification after accounting for diagnostic GGG and AI-identified cribriform morphology. Additionally, AI-GUR provided information that was not strongly correlated with prognostic risk methodologies (CAPRA, Prolaris). Thus, AI-GUR has potential utility above and beyond what is currently available to aid decisions regarding AS for disease management.

Whereas standard clinical risk variables such as GGG and NCCN risk group did not provide significant information to AI-GUR, PSA at diagnosis provided marginally significant (p=0.04) information. This relationship should be further explored, especially given the weight PSA has in the CANARY PASS Calculator[9].

Cribriform morphology is definitionally absent from GGG1 and increases in prevalence as GGG increases. In the ProtecT study[10], cribriform morphology was identified in ∼13% of the pathologically re-reviewed cohort, and evidence of cribriform positivity was associated with significantly higher risk of developing metastases[11]. As demonstrated herein, the AI-GUR predictive model differentiates risk of GGG reclassification events independent of the cribriform sub-feature of the PATHOMIQ_PRAD algorithm.

Further validation in independent cohorts is ongoing to determine the impact of biopsy modality (systematic, image-guided) on the performance of AI-GUR. If AI-GUR was predicting true disease progression, the distribution of upgrade events would be enriched at later time points, which was not observed in first follow-up biopsies as far as 5-years post-diagnosis (Supplemental Figure 2). Although the model does not directly distinguish disease progression from biopsy-sampling variability (due to factors such as tumor heterogeneity), that distinction would not impact clinical decisions as currently understood: a confirmatory biopsy has importance irrespective of the cause of reclassification. DPAI-enabled models, such as AI-GUR, have promising potential as decision aids in the management of PCa, and future work will evaluate its utility in real-world scenarios.

## Supporting information

TRIPOD-AI Checklist

## Acknowledgements

The authors would like to thank Sabra Powell, Ken Eyring, and Dana Case for lab logistics support; Lori Windorski for product support; Amanda Steele and Brooke Hullinger for medical writing support; Raman Randwawa, Parag Jain, Devraj Mondal, and Murali Selvrajan for their contributions to the development of PRAD.

## Data availability statement

Due to the nature of the research, due to ethical/legal/commercial reasons, supporting data is not available.

## Reference Annotations

(2) of considerable interest: The NCCN Clinical Practice Guidelines in Oncology (NCCN Guidelines®) are cited to ground active surveillance candidacy/confirmatory biopsy context and the clinical framework motivating this work.

(1) of interest: Awamlh et al. describe use of active surveillance vs definitive treatment in low- and favorable intermediate-risk prostate cancer, supporting the manuscript’s clinical context for AS utilization.

(3) of considerable interest: Lin et al. address identification of men with low-risk biopsy-confirmed prostate cancer as candidates for active surveillance in the context of Prolaris results, aligning with the manuscript’s discussion of AS eligibility and risk stratification.

(7) of considerable interest: Cuzick et al. report on the prognostic value of a cell cycle progression signature (Prolaris) for prostate cancer outcomes in a conservatively managed biopsy cohort, providing key background for molecular prognostic testing referenced in the manuscript.

(4) of interest: Huang et al. present an AI-powered method for prediction of prostate cancer recurrence after prostatectomy, serving as relevant precedent for AI approaches applied to prostate cancer risk prediction.

(6) of interest: Fay et al. describe an AI-based digital histologic classifier with independent blinded validation, supporting broader context for validation expectations in AI pathology classifiers.

(10)of considerable interest: Hamdy et al. report fifteen-year outcomes after monitoring, surgery, or radiotherapy for prostate cancer, providing long-term management context relevant to AS as a strategy.

(11)of interest: Sushentsev et al. focus on cribriform-positive vs cribriform-negative prostate cancer across active monitoring, surgery, and radiotherapy, aligning with the manuscript’s emphasis on cribriform morphology as an important pathological feature.

