## Supplemental Material for "Development and validation of a digital pathology artificial intelligence (DPAI)-based biomarker predicting risk of Gleason grade group reclassification for patients who are candidates for active surveillance"

2-step Implementation of the PATHOMIQ_PRAD Algorithm

First, slides are reviewed by a quality control algorithm that also detects regions of tissue without scanning and staining problems, or other artifacts. The algorithm uses a pretrained ResNet-18 model to classify patches as ink or tissue and generates tissue masks from the LAB A-channel using an adaptive threshold with morphological refinement. Slide quality is assessed using focus (Laplacian variance), noise (small connected-component fraction), fragmentation (connected component density), and staining-contrast metric (A-channel standard deviation). Low-quality slides revert to Otsu-based masking, and slides with insufficient patches are excluded from downstream inference.

Second, a series of multi-resolution convolutional neural networks (triple Inception v1 with shared weights) are used to predict localizations of cancer and the Gleason grades found throughout the epithelial regions. Nuclei are detected using a watershed method and then patches of size 64x64 px (0.25 mpp), 256x256 (0.25 mpp), and 128x128 (1.0 mpp) are extracted around points centered on nuclei. Specialized cancer detection and cancer grading models are used for inference, assigning each multi-resolution image patch a label indicating if cancer is found in the image patch, and if so, what Gleason grade is identified. These metrics are then summarized, resulting in total tissue volume, tumor volume, percent Gleason pattern (GP)3, percent GP4, percent GP5, percent cribriform, and perineural invasion presence.

**Supplemental Figure 1:** Patient flow diagram. Boxes outlined in blue indicate analysis sets.

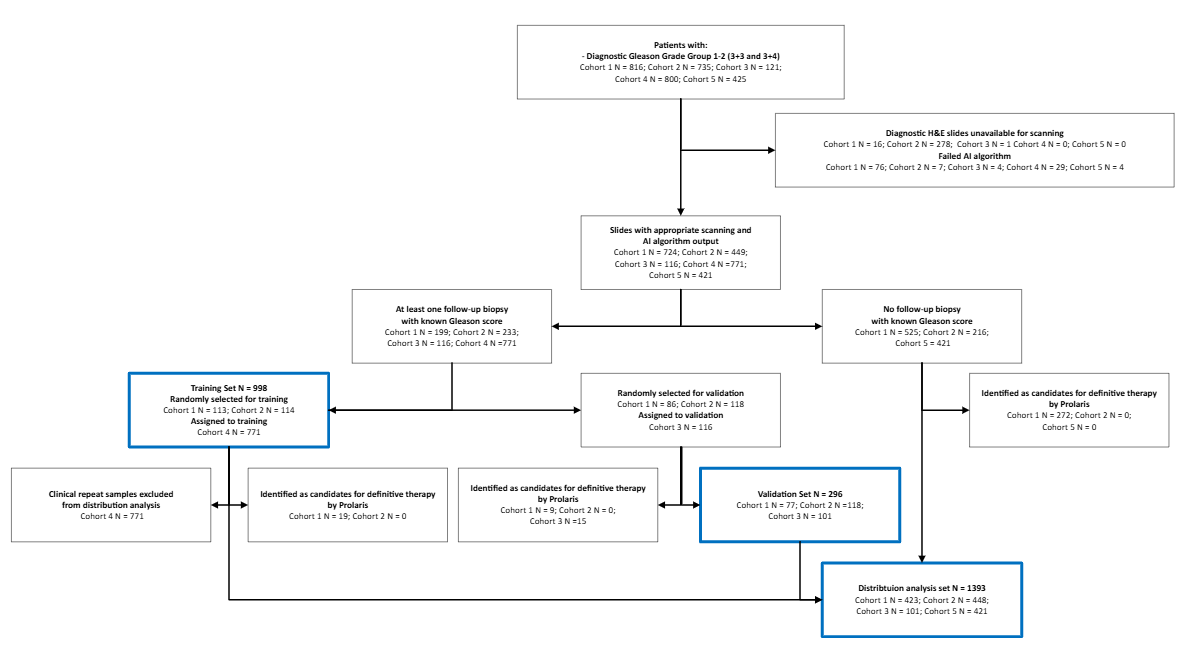

**Supplemental Figure 2:** Distribution of time between diagnosis and first follow-up biopsy in the validation cohort, colored by Gleason upgrade status.

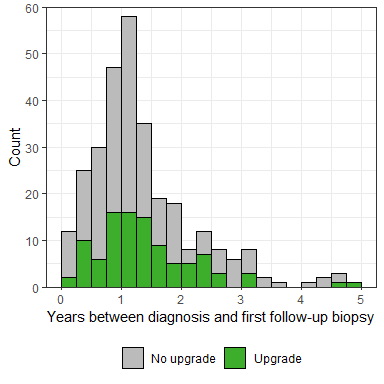

**Supplemental Figure 3:** Distribution of extent of Gleason pattern 4 cribriform morphology identified as a portion of (A) AI-identified tissue or (B) AI-identified cancer, separated by Gleason upgrade status.

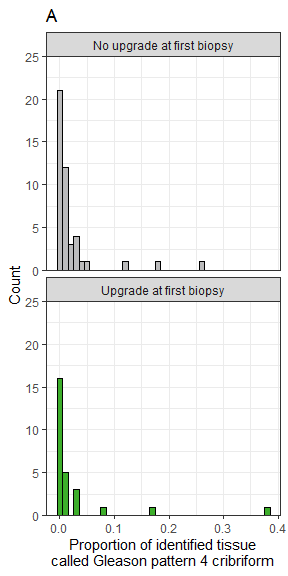

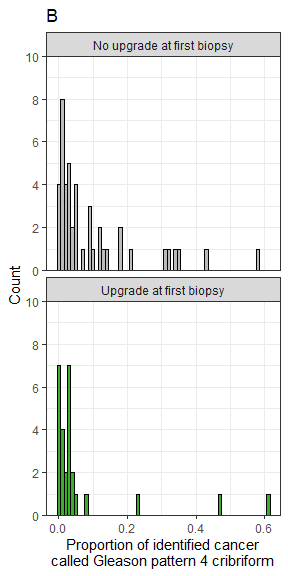

**Supplemental Table 1:** Characteristics of the training cohort, N = 998.

|  | N (%) or Median (IQR) |
| --- | --- |
| Diagnostic Gleason Score |  |
| 3+3 | 590 (59.1%) |
| 3+4 | 408 (40.9%) |
| NCCN Risk Category* |  |
| Low | 565 (56.6%) |
| Favorable Intermediate | 384 (38.5%) |
| Unfavorable Intermediate | 48 (4.8%) |
| High | 0 (0%) |
| Years to first follow-up biopsy | 1.1 (0.9, 1.8) |
| Upgrade at first follow-up biopsy | 474 (47.5% |

* One patient has missing percent positive cores and so NCCN Favorable vs Unfavorable Intermediate risk cannot be distinguished.
